# Explanations for higher-than-expected mortality from April 2021: a scoping review

**DOI:** 10.1101/2024.05.02.24306706

**Authors:** Fiona Scott, Gerry McCartney, David Walsh, Sarah Wild, David Rae, Julie Ramsay, Grant Donaghy, Margaret Douglas

## Abstract

**Objectives:** Many countries have continued to experience a higher-than-expected number of deaths following the peaks in mortality observed in the first year of the Covid-19 pandemic. This scoping review aims to identify the different explanations proposed for sustained higher-than-expected mortality beyond the first pandemic year.

**Study design:** Scoping review

**Methods:** A systematic search of databases and grey literature sources was completed to identify English-language records proposing or investigating hypotheses for higher-than-expected mortality from April 2021 onwards in (sub)populations of high-income countries. Papers focused on survival following a diagnosis or intervention were excluded. Results were summarised narratively, and existing research prioritisation frameworks were adapted and applied to identify the hypotheses proposed as highest priority for further research.

**Results:** Seventy eligible papers were identified. Most were opinion pieces or simply presented trends; few included investigation of suggested hypotheses. Numerous explanations for higher-than-expected mortality were proposed, with hypotheses relating to direct Covid-19 mortality, sequalae of Covid-19 infection, the health service impacts of the pandemic, wider pandemic impacts and socioeconomic factors identified as highest-priority for further research.

**Conclusions:** The causes of continued higher-than-expected mortality are likely to be multiple and potentially interactive. This review will help to shape research into current mortality trends, with a critical understanding of this topic essential for achieving evidence-informed policy.

## Introduction

Following the high levels of mortality seen globally during the peak waves of Covid-19 in 2020 and early 2021, many countries, including the UK, have continued to experience mortality levels higher than in previous years^1^. Although effective vaccines have been available since December 2020, Covid-19 remains an important cause of death and accounts for some of this continuing excess mortality. The number of non-covid deaths also remains high^2^ ^3^. The people most susceptible to dying from Covid-19 in 2020/21 included many who would otherwise have died within the following few years, resulting in a degree of mortality displacement ^4^. This would be expected to lead to period of lower mortality compared to previous years, so the continuing high rates are unexpected.

Different metrics can be used to measure mortality trends. Excess mortality is a measure that describes seasonal differences by comparing observed mortality data (sometimes crude death counts, sometimes crude, non-standardised, mortality rates per unit population) with the average mortality for the same time week or month in previous years^5^. This often uses the five previous years as baseline, excluding 2020 to remove the effect of the first Covid-19 waves of mortality ^6^. Over longer time periods, changes in the age structure of the population (including ageing and age-selective migration) confound crude mortality counts and rates, making them less reliable as a monitoring tool^1^. Using age-standardised mortality rates or life expectancy avoids this, but these are generally not available as quickly or published as frequently as crude measures^7^. Recent analysis of UK data show that, while using age-standardised mortality rates provides more accurate estimates of excess deaths, there were still substantially high levels of excess deaths between 2020 and 2022 even when this was accounted for.^8^

This recent concern about higher-than-expected mortality comes on top of an unprecedented change in the longer-term trend. After decades of continuous improvements, average mortality rates across the UK population stopped improving after around 2012 and mortality rates in people living in more deprived populations actually started to increase^9, 10^. Other high-income countries experienced similar trends, and the evidence shows that this has been caused by austerity policies introduced widely from around 2010 onwards^11,12^ The wider effects of the pandemic control measures (including social, economic, and healthcare disruption) and the recent rise in inflation with its impact on real incomes, have exacerbated these impacts ^13,14^. However, the full range of causes of the high mortality after the peak waves of the pandemic remains somewhat unclear and contested.

In order to inform research into this mortality phenomenon, we aimed to identify and assess proposed hypotheses for the higher-than-expected mortality in high income countries observed beyond the first year of the Covid-19 pandemic.

## Methods

Our research question was: what explanations have been proposed for higher-than-expected mortality in high income countries persisting beyond the first year of the Covid-19 pandemic, and what empirical evidence is available either supporting or refuting these hypotheses? We chose this time cut-off to exclude the peak waves of mortality caused by Covid-19 disease before effective vaccines were available. In the UK and many other countries peak mortality from Covid-19 occurred in April and May 2020, and November 2020 to February 2021.

We used JBI methodology for scoping reviews^15^ to inform the study and published a protocol for the review on MedRxiv^16^.

### Search strategy

We searched for academic studies on Medline, Embase and Google Scholar and grey literature on websites including government and public health sites, and by using Google. Our search strategy used ‘excess mortality’ ‘life expectancy’ and ‘increased death’, together with a search string designed to exclude low-income settings and was piloted on Medline. We restricted the search to studies published in English from 2021 onwards. The search strategy is detailed in the review protocol pre-print.^16^

### Screening and selection of studies

We imported search results into Covidence for screening and review against eligibility criteria. Inclusion criteria were: papers that proposed theories or evidence to explain worse-than-expected mortality or life expectancy; in high income countries; from April 2021 onwards compared to pre-pandemic levels; all-cause and cause-specific mortality; in the whole population or any specific sub-populations. We excluded: studies of mortality or survival following a healthcare intervention (due to our focus on mortality trends rather than treatment efficacies); studies comparing survival in people with and without a diagnosis; and studies from countries that have not experienced worse than expected mortality from April 2021.

All authors took part in screening and review. Two reviewers independently reviewed all titles and abstracts, with disagreements discussed in meetings to achieve consensus. Similarly, two reviewers completed full text reviews and resolved differences by consensus.

### Data extraction and management

We developed a template on Covidence to extract data on: author, dates of mortality trends considered, setting, mortality metrics, definition of baseline mortality, population(s), causes, hypotheses proposed, paper type, study type, and findings (if relevant). Two team members extracted data from each paper independently, with differences resolved by consensus.

### Synthesis

We grouped papers according to the proposed hypotheses and noted empirical evidence provided in support of each. We then summarised the hypotheses narratively. We did not critically appraise the papers because the main aim was to identify hypotheses and we found few analytical studies investigating these.

### Prioritisation

In order to prioritise hypotheses for further work, we identified relevant criteria from an existing review of research prioritisation frameworks^17^ and adapted these to focus on two key questions:

1. Is there sufficient existing research or research effort on this hypothesis?
2. Would additional research on this area have potential to change policy or practice?

Due to potential variation in the relevance of each hypothesis to mortality trends across different high income countries, we chose to focus on a UK context when completing this prioritisation exercise. Based on discussion amongst authors, we ranked hypotheses highest priority where we judged that there was insufficient existing research or research effort likely to be applicable to a UK setting, and where additional research was felt to have the potential for changing policy or practice in the UK. This ranking was based on our own views and opinions, and we did not seek external input into the prioritisation exercise.

## Results

### Description of included studies

We identified 3,969 papers through database searching, and 437 from grey literature sources (435 records) and citation tracking (2 records). We removed 1,605 duplicate references and excluded 2,641 at title and abstract screening, leaving 160 papers for full-text review. At full text review we excluded 90 papers which did not meet eligibility criteria, leaving 70 included papers (Figure 1). ^1, 18–86^

**Figure 1:**
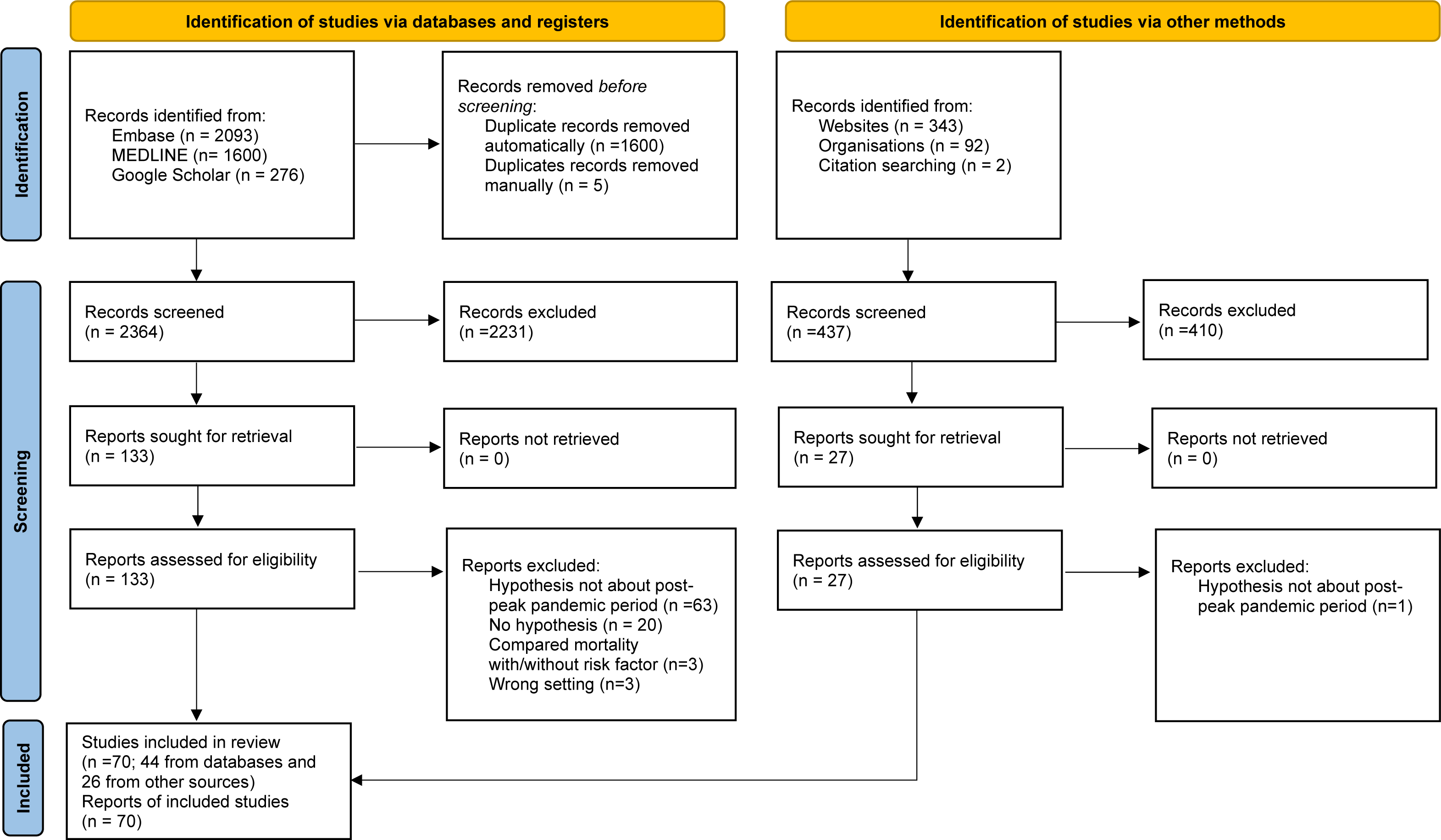
PRISMA 2020 flow diagram for systematic reviews including searches of databases, registers, and other sources. Figure adapted from the PRISMA 2020 statement (Page, McKenzie, Bossuyt et al., 2021) ^87^

Although we had only included papers that considered mortality beyond April 2021, many studies included mortality data from 2020 but did not always differentiate between time periods in their analyses. The dates of mortality trends considered across all papers are shown in Figure 2. Other characteristics of papers are presented in Table 1.

**Figure 2:**
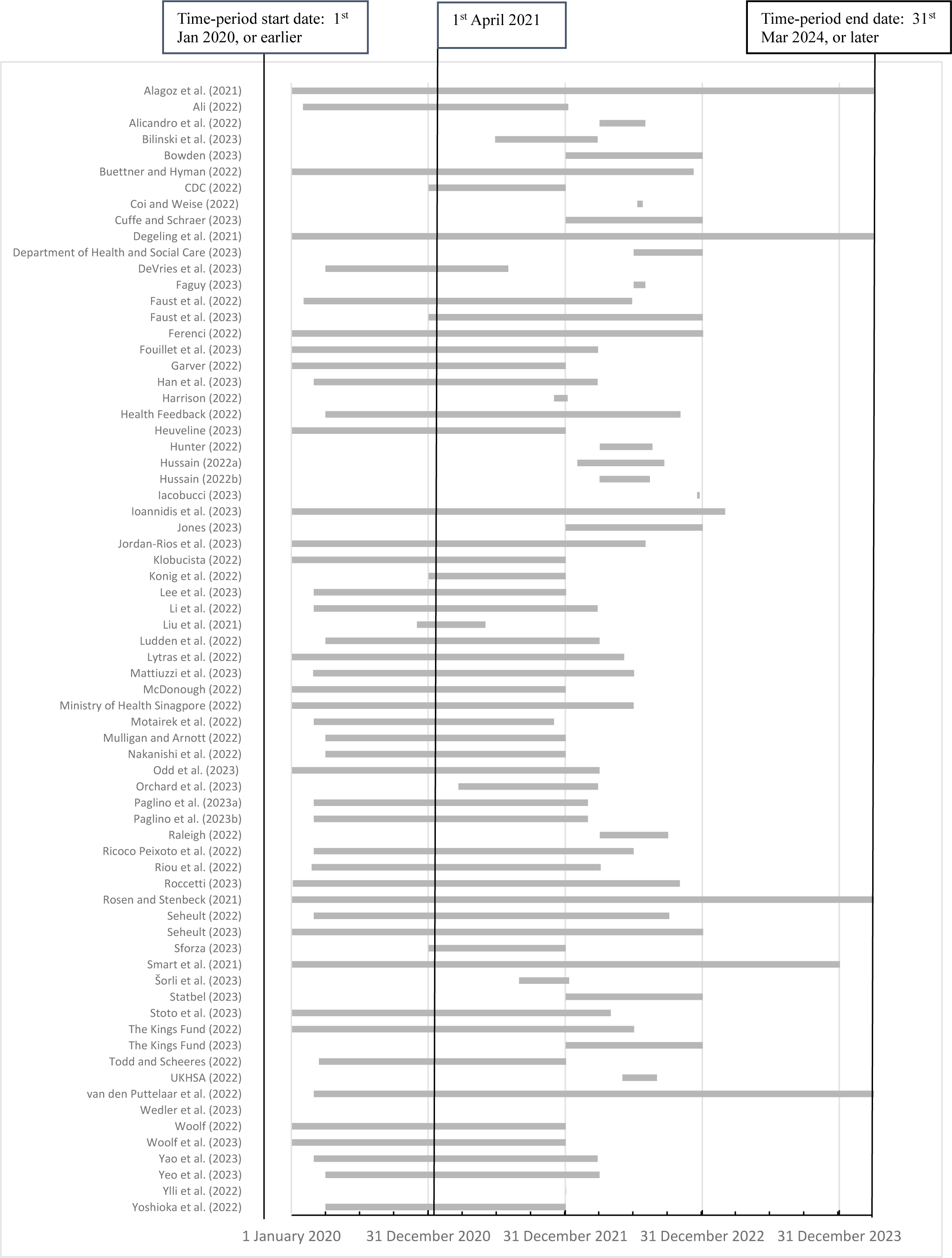
Dates of mortality trends considered across included papers, between 01 January 2020 (or before) – 31 December 2023 (or after). *\**Mortality forecasting study. *Note: the time periods of mortality trends considered by Wedler et al.* ^80^ *are not captured by the above chart (paper projects mortality from 2026)*.

**Table 1:**
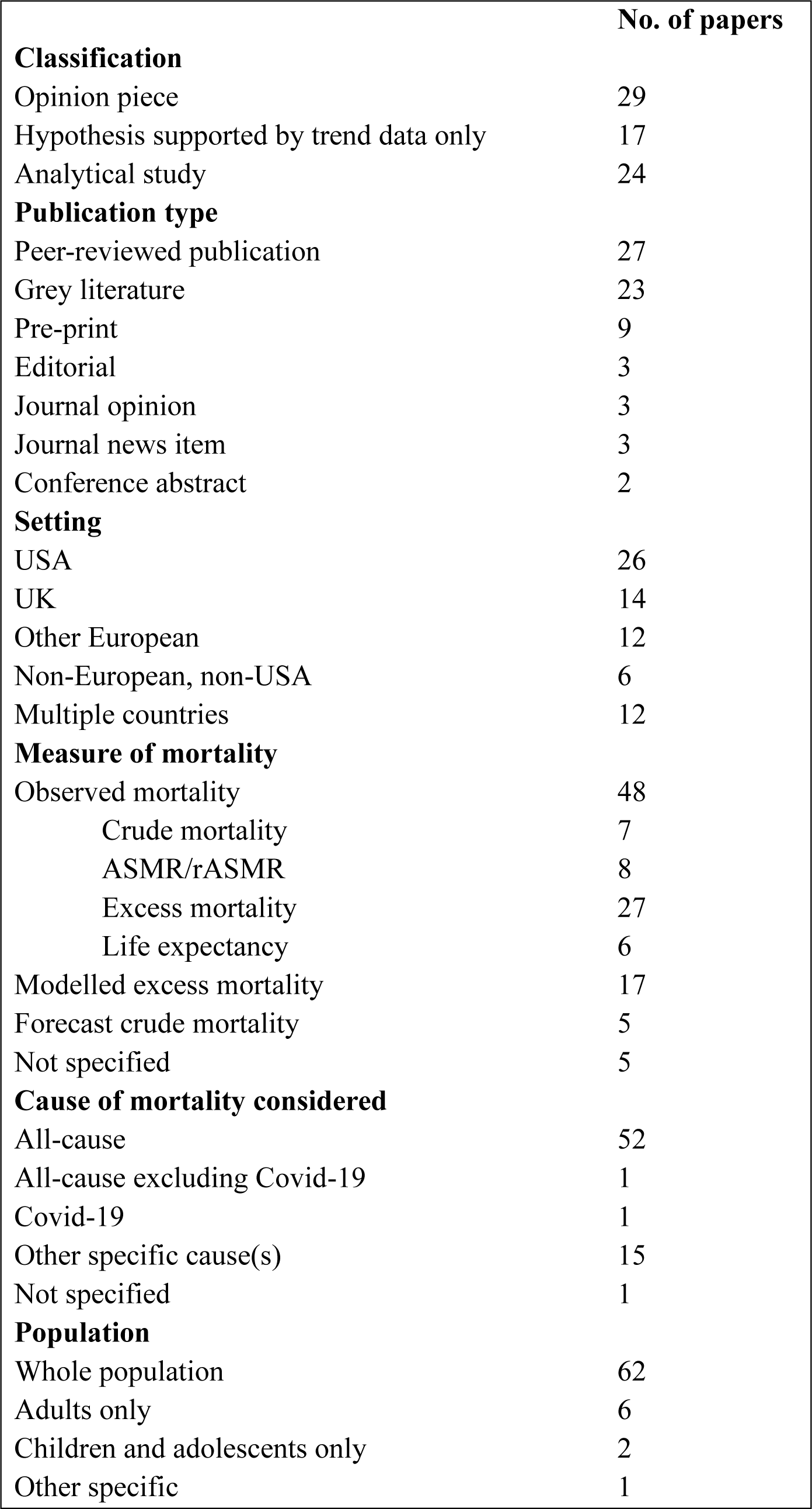
Characteristics of included papers. ASMR = Age-standardised mortality rates; rASMR = relative age-standardised mortality rates*. Note: Categorisations of papers may not add up to the total number of included papers if, e.g., multiple measures or causes of mortality were considered*.

### Hypotheses identified

Papers identified a range of hypotheses and pathways to explain continued higher than expected mortality as shown in Table 2. The most frequently proposed explanations related to direct Covid-19 mortality (35 papers), health service impacts (34 papers) and other pandemic impacts (18 papers). Fifty-eight papers suggested more than one hypothesis.

**Table 2:** Hypotheses and pathways explaining continued higher than expected mortality.

| **Hypotheses** | **No. of studies** | **Pathways** | **Analytical studies of hypotheses** |
| --- | --- | --- | --- |
| Direct Covid-19 | 35 | Covid-19 disease still accounting for acute deaths<br>Underestimation of cases due to undetected infections<br>Continuing direct Covid-19 deaths due to:<br>• Low vaccine uptake<br>• Vaccine hesitancy<br>• Poor implementation of NPIs<br>Underpinned by:<br>• Poor targeting of vaccination<br>• Misinformation affecting controls<br>• Poor PH infrastructure | 6 |
| Sequelae of Covid-19 | 14 | Mortality associated with Post Covid-19 Condition.<br>Higher CV risk following SARS CoV2 infection<br>Long term CNS damage following SARS CoV2 infection | 1 |
| Health service impacts of pandemic | 34 | Longer term impacts of paused healthcare during the peak pandemic waves causing delayed screening, care or preventive interventions<br>Continued healthcare disruption due to<br>• Backlog of patients<br>• Staff absences from burnout or continuing Covid-19 policies<br>• Damaged medical education<br>Change in health seeking behaviour due to fear of transmission or earlier communications or experiences discouraging attendance | 3 |
| Wider impacts of pandemic | 19 | Pandemic related unemployment<br>Effects of social distancing on health-related behaviours:<br>• Sedentary behaviour<br>• Unhealthy diet<br>• Increased substance use<br>Social isolation causing sleep disturbance and declined cognitive function<br>Stress and distress causing suicidal ideation and myocardial ischaemia<br>Caregivers' isolation, distress or illness affecting those cared for<br>Increased interpersonal violence<br>Increased traffic speeds | 2 |
| Influenza and other infections | 9 | Suppressed influenza and other infections during social distancing, leading to lowered immunity and then resurgence once restrictions eased | 0 |
| Vaccine- associated deaths | 5 | Vaccination causes mortality | 3 |
| Other socioeconomic causes not necessarily related to pandemic | 7 | Poverty and cost of living crisis<br>Social vulnerability – increased mortality pre-pandemic, increased susceptibility to Covid-19, continuing to increase mortality | 2 |
| High prevalence of chronic conditions | 7 | Previously high prevalence of chronic conditions exacerbating Covid-19 and continuing to cause high mortality<br>Underpinned by unhealthy environments, poverty and inequality, ultra-processed foods<br>Exacerbated further by social distancing | 0 |
| Wider societal causes | 7 | Gun violence<br>Racism<br>Conflict<br>MH crisis<br>Opioid crisis<br>Poor rural infrastructure<br>Lack of UHC<br>Cuts to PH budgets | 0 |
| Extreme weather temperatures | 12 | Heatwaves during summer 2022<br>Cold snap in December 2022 | 1 |
| Displacement | 9 | Repressed pathogens affecting baseline during restrictions<br>Lower mortality in later periods following pandemic excess<br>Higher mortality in late 2022 due to lower-than-expected winter deaths in 2021/22 | 0 |
| Artefact | 2 | High excess mortality measure reflects aging population<br>High excess mortality measure in Germany from effect of random fluctuation on baseline used in WHO spline model | 1 |

### Papers including analysis of hypotheses

A minority of the papers included analyses aimed at testing the specified hypotheses for high mortality. Twenty-nine of the papers were opinion pieces, commenting on mortality data published elsewhere. Seventeen presented estimates of all cause and/or cause specific excess mortality and then suggested reasons for the observed trends. Twenty-four were analytical studies of various designs including ecological, time series and prospective modelling studies. Two of the ecological studies compared mortality between states or countries and suggested reasons explaining high mortality settings without investigating these further.^21^ ^74^ We identified no analytical studies that investigated the importance of influenza or other infections, prevalence of chronic conditions, wider societal issues or displacement as hypotheses for continuing high mortality.

Six studies investigated direct Covid-19 as a cause of excess mortality. Two estimated unascertained Covid-19 mortality but both presented estimates for cumulative mortality from 2020 to early 2022, so may be less relevant in later time periods.^65^ ^49^ Two Italian time series studies using different mortality metrics reached opposing conclusions about the association between Covid-19 and all-cause mortality in 2022.^54^ ^66^ Two ecological studies correlated excess mortality and vaccination rates. One studied cumulative mortality from 2020 to January 2022 in 50 countries and found higher mortality in countries with lower vaccination and poor enforcement.^85^ The other found an inverse correlation between mortality and vaccination in US states in 2021 but not in 2022.^32^

The only analysis of sequelae of Covid-19 was a cohort study that found higher mortality at one year in people who had had a Post-Covid-19-Condition in 2020, compared with matched controls. This study used a claims-based definition of post-Covid-19 condition, based on the recording of diagnoses consistent with Covid-19 related symptoms in the 5-12 weeks following infection ^29^

We found five prospective modelling studies that estimated future mortality relating to one of the identified hypotheses. These included three studies that estimated future cancer deaths resulting from delayed screening, diagnosis or treatment when non-covid health services were paused.^18^ ^27^ ^79^ One study modelled the impact of increased unemployment in Sweden in April 2020 on mortality for the next 9 years.^67^ The final modelling study estimated mortality from heatwaves in the Eastern Mediterranean up to 2100. ^80^

A study of the correlation between suicide in Japan with unemployment and eating out (as a proxy for isolation) found these did not explain the trends seen.^59^

Two ecological studies correlated mortality with social vulnerability or economic indicators. Both report a positive correlation, but one considered cumulative mortality from 2020 ^44^ and the other presented data for all of 2021, which had a lower correlation than 2020.^57^

In addition to the ecological studies noted above that found an inverse correlation between mortality and vaccination rates ^32,85^, three other studies investigated vaccine related deaths, but with conflicting findings.^51,53,72^ However, all three of these have methodological limitations that mean findings should be treated with caution.

Finally, one study critiqued excess mortality measures in Germany and showed excess mortality to be lower when an alternative method was used to assess baseline mortality. ^33^

Table 3 shows the application of the prioritisation criteria to the identified hypotheses. With reference to a UK setting, five hypotheses are identified as high priority (direct Covid-19, sequelae of Covid-19, health service impacts of the pandemic, wider impacts of the pandemic, and other socioeconomic causes), two as medium priority (wider societal impacts and displacement), and five as low priority (high prevalence of chronic conditions, influenza and other infections, vaccine-associated deaths, extreme weather, and artefact).

**Table 3:**
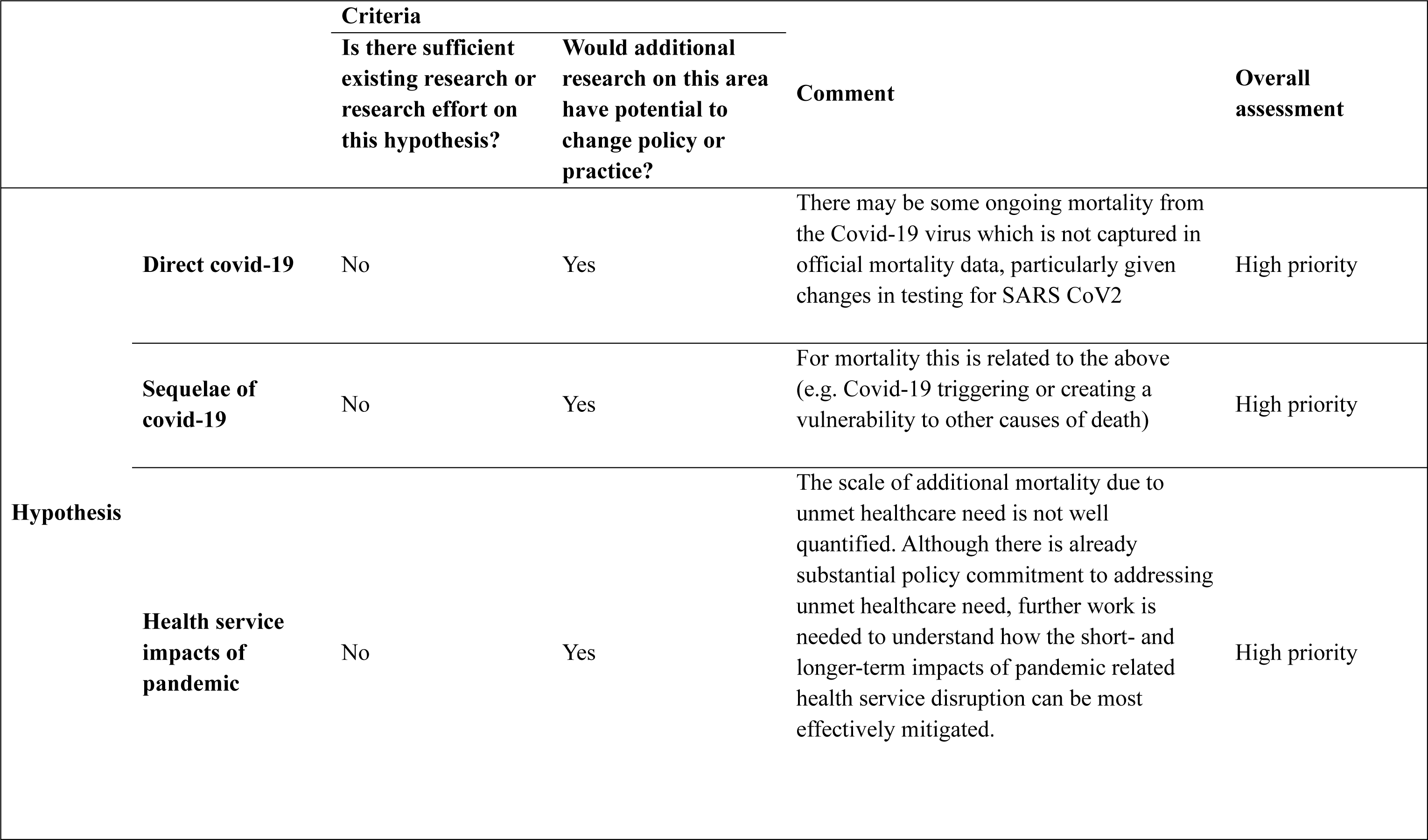
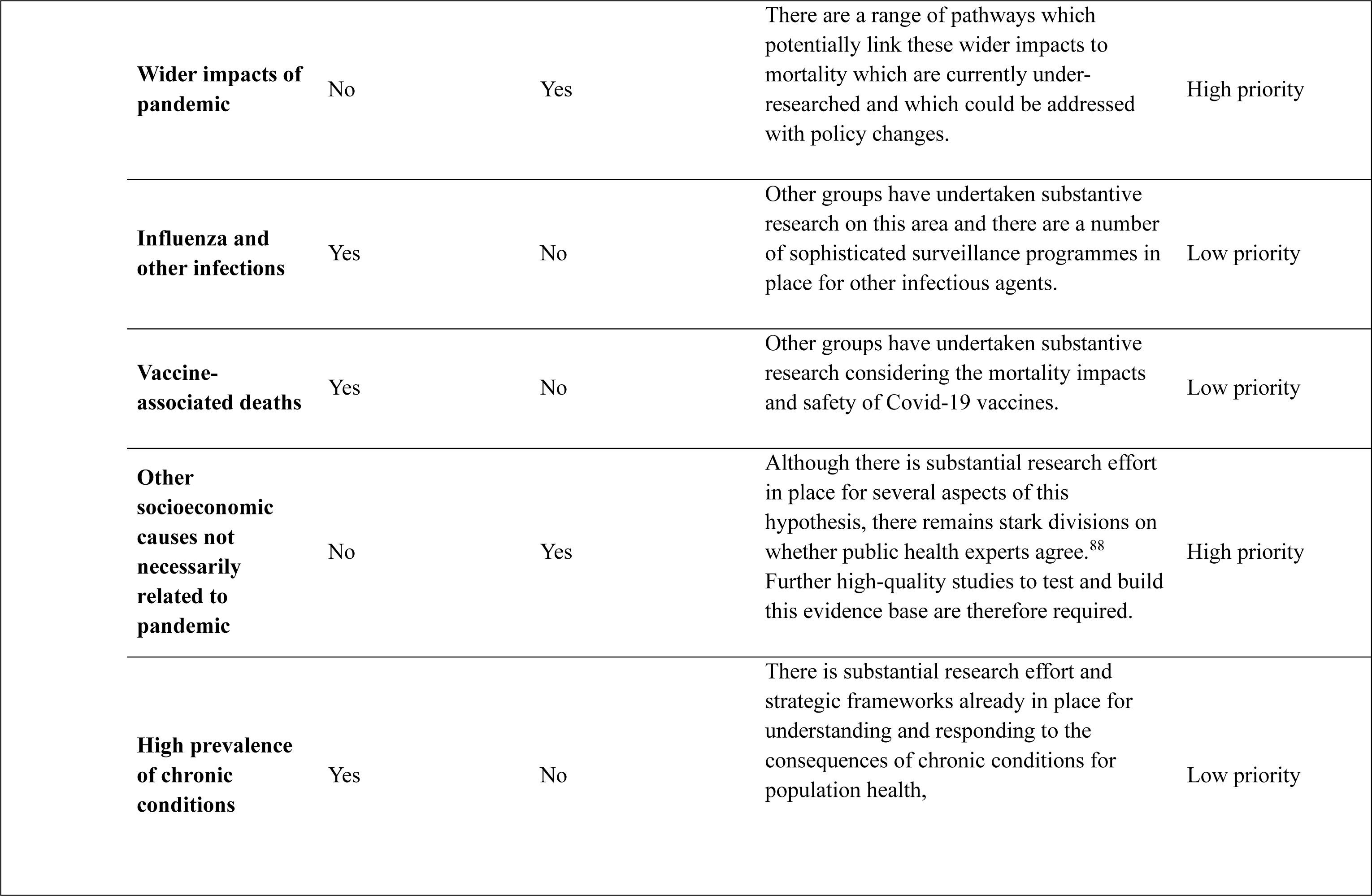
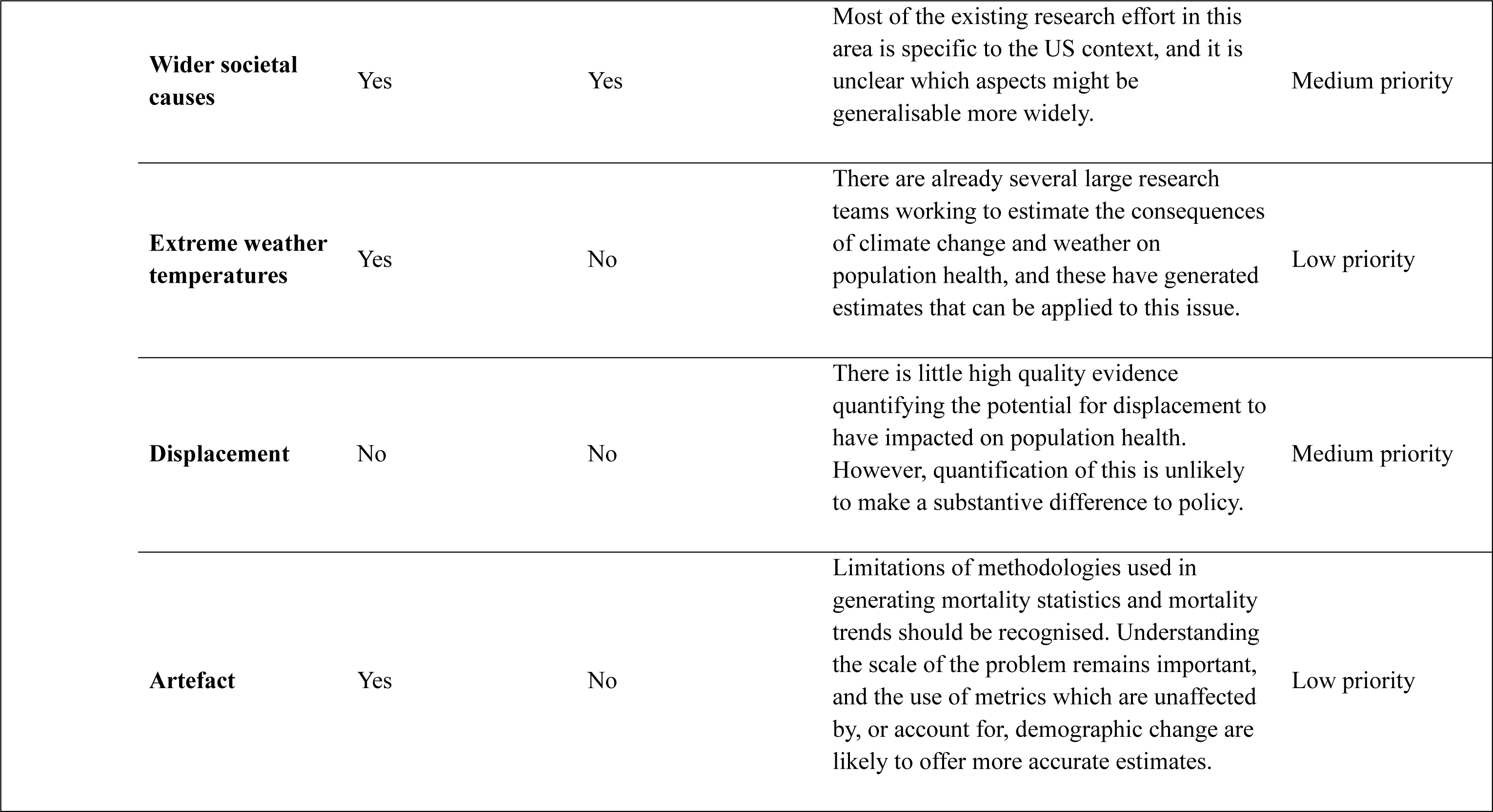
Prioritisation of hypotheses for future research.

|  |  | **Criteria** |  | **Comment** | **Overall assessment** |
| --- | --- | --- | --- | --- | --- |
|  |  | **Is there sufficient existing research or research effort on this hypothesis?** | **Would additional research on this area have potential to change policy or practice?** |  |  |
| **Hypothesis** | **Direct covid-19** | No | Yes | There may be some ongoing mortality from the Covid-19 virus which is not captured in official mortality data, particularly given changes in testing for SARS CoV2 | High priority |
|  | **Sequelae of covid-19** | No | Yes | For mortality this is related to the above (e.g. Covid-19 triggering or creating a vulnerability to other causes of death) | High priority |
|  | **Health service impacts of pandemic** | No | Yes | The scale of additional mortality due to unmet healthcare need is not well quantified. Although there is already substantial policy commitment to addressing unmet healthcare need, further work is needed to understand how the short- and longer-term impacts of pandemic related health service disruption can be most effectively mitigated. | High priority |

|  |  |  |  |  |
| --- | --- | --- | --- | --- |
| **Wider impacts of pandemic** | No | Yes | There are a range of pathways which potentially link these wider impacts to mortality which are currently under-researched and which could be addressed with policy changes. | High priority |
| **Influenza and other infections** | Yes | No | Other groups have undertaken substantive research on this area and there are a number of sophisticated surveillance programmes in place for other infectious agents. | Low priority |
| **Vaccine-associated deaths** | Yes | No | Other groups have undertaken substantive research considering the mortality impacts and safety of Covid-19 vaccines. | Low priority |
| **Other socioeconomic causes not necessarily related to pandemic** | No | Yes | Although there is substantial research effort in place for several aspects of this hypothesis, there remains stark divisions on whether public health experts agree.<sup>88</sup> Further high-quality studies to test and build this evidence base are therefore required. | High priority |
| **High prevalence of chronic conditions** | Yes | No | There is substantial research effort and strategic frameworks already in place for understanding and responding to the consequences of chronic conditions for population health, | Low priority |
| **Wider societal causes** | Yes | Yes | Most of the existing research effort in this area is specific to the US context, and it is unclear which aspects might be generalisable more widely. | Medium priority |
| **Extreme weather temperatures** | Yes | No | There are already several large research teams working to estimate the consequences of climate change and weather on population health, and these have generated estimates that can be applied to this issue. | Low priority |
| **Displacement** | No | No | There is little high quality evidence quantifying the potential for displacement to have impacted on population health. However, quantification of this is unlikely to make a substantive difference to policy. | Medium priority |
| **Artefact** | Yes | No | Limitations of methodologies used in generating mortality statistics and mortality trends should be recognised. Understanding the scale of the problem remains important, and the use of metrics which are unaffected by, or account for, demographic change are likely to offer more accurate estimates. | Low priority |

## Discussion

This scoping review identified a range of different proposed explanations for the higher-than-expected mortality rates observed in multiple high-income countries persisting beyond the first year of the Covid-19 pandemic. While some hypotheses were tested using empirical data, those were in the minority; several papers presented analyses of trends, but most did not go beyond discussion of potential or likely causal factors. The most commonly proposed explanations related to the direct and indirect effects of the Covid-19 pandemic, including: continued significant mortality from Covid-19 itself; adverse longer-term effects of SARS-CoV-2 infection (including higher mortality risk); the negative impact of the pandemic on quality of, and access to, healthcare; and wider societal effects such as social isolation, increased poverty and health-harming behavioural responses. Non-Covid explanations that were suggested included extreme weather events, and political and/or societal factors such as funding cuts, drug use, social vulnerability and increased poverty levels. The different proposed explanations are not mutually exclusive; it seems likely that the excess will be best explained by a combination of multiple, inter-related, factors.

The implications of the work are obvious and important: we need to attempt to understand the precise causes of the higher mortality to be able to address them. It is important not to base policy on unevidenced theoretical assertions, as has already happened in the UK^89^. There is therefore a need for further research to investigate some of the proposed explanations summarised here. We have identified the explanations that are highest priority for this further research.

### Strengths and weaknesses

To our knowledge, this is the first scoping review of this important topic. The review process has been systematic, based on independent screening, review and data extraction, and the methods are transparent and replicable. Searches included a considerable amount of grey literature which is important for the subject matter.

However, a number of limitations are acknowledged. As the primary objective was to identify all relevant hypotheses (assessments and testing of which could form the basis of future work), we did not undertake a critical appraisal of the many different studies that emerged from the literature searches. The available literature was limited by the short time period included. The definition of the time period was also problematic, as discussed further below.

In relation to our prioritisation of hypotheses for further research relevant to a UK context, we prioritised the hypotheses but have not prioritised specific research proposals to investigate these. Specific research proposals will then also need to be prioritised in relation to their scale, data availability, likely impact, robustness and other characteristics. The application of prioritisation criteria was based on author opinion only and we were unable to achieve unanimous consensuses for all hypotheses considered. We recognise that there are likely to be varied views on the prioritisation of hypotheses within the public health community.

### Comparison with other studies

In the UK, and in a number of other high-income countries such as the USA, high levels of excess mortality had already been highlighted prior to the pandemic^10^ ^90^. This has been principally attributed to economic ‘austerity’ policies which have had particularly adverse effects on poorer populations, resulting in increasing death rates and subsequent widening of inequalities. ^11,91^. The extent to which the post-pandemic high rates of mortality largely reflect those pre-pandemic trends (albeit exacerbated by Covid-19) did not feature prominently in the literature, yet it is likely to be important (and is the subject of ongoing research)^92^. This is also relevant to the debated issue of how ‘excess deaths’ are defined, including the use of a five-year comparative baseline. For example, mortality rates in the UK in 2022 may have been somewhat higher than those of 2017; but they were dramatically higher compared to predicted trends based on pre-austerity data (prior to 2010s)^93^. This is highlighted by new guidance on measuring excess deaths in the UK which suggests that by 2023, the excess compared to the recent five-year baseline was minimal ^8^; however, a longer-term perspective (based on projecting forward pre-austerity trends) paints a different picture.

The continuing impact of Covid-19 featured prominently in the results of the literature searches, as perhaps might have been expected. This is clearly also relevant to the issue of how the ‘peak’ Covid and post-peak Covid periods are defined. In terms of direct Covid-19 deaths, analyses of Scottish data are informative: in 2020 and 2021, an estimated 12,751 and 10,651 deaths respectively were recorded as involving Covid-19, representing age-standardised rates of 128 and 108 per 100,000 population. However, although the equivalent figures for 2022 were considerably lower, at 6,247 deaths (an age-standardised rate of 74 per 100,000 population), they were still high^94^. The wider effects of Covid on healthcare have been much documented^95^, while broader societal impacts of the pandemic and its associated control measures were assessed and predicted in a number of reviews^96,97^. It seems likely that a combination of pre-pandemic trends and these direct and indirect effects of Covid-19, alongside other factors such high inflation (the ‘cost of living crisis’) – which has been associated with increased death rates in the population^13^ – will be important in explaining the current higher levels of mortality. However, further research is urgently needed to quantify the contribution of each, and of other potential factors, to inform the most effective policy response.

## Data Availability

All data produced in the present study are available upon reasonable request to the authors.

## Author Contribution Statement

The study was conceived by MD. The research questions and analysis plan were agreed by all authors. GD designed the search strategy and performed the searches. All other authors participated in screening, selection, data extraction and interpretation. MD, GM and FS drafted the manuscript. All authors provided substantial critical input to improve the manuscript and all authors approved the final draft.

## Ethical approval

This paper is a scoping review; ethical approvals were not required.

## References

1 Raleigh V. What is driving excess deaths in England and Wales? BMJ 2022; 379:o2524 10.1136/bmj.o2524.

2 Office for Health Improvement and Disparities. Excess mortality in England, https://app.powerbi.com/view?r=eyJrIjoiYmUwNmFhMjYtNGZhYS00NDk2LWFlMTAtOTg0OGNhNmFiNGM0IiwidCI6ImVlNGUxNDk5LTRhMzUtNGIyZS1hZDQ3LTVmM2NmOWRlODY2NiIsImMiOjh9; 2024 [last accessed 8 April 2024].

3 National Records of Scotland. Monthly Mortality Analysis, Scotland, March 2023, https://www.nrscotland.gov.uk/files//statistics/vital-events/monthly-mortality/monthly-mortality-march-23-report.pdf; 2023 [last accessed 8 April 2024].

4 Islam N, Shkolnikov V M, Acosta R J, Klimkin I, Kawachi I, Irizarry R A, et al. Excess deaths associated with covid-19 pandemic in 2020: age and sex disaggregated time series analysis in 29 high income countries. BMJ 2021; 373 :n1137 10.1136/bmj.n1137.

5 Office for National Statistics. Comparing different international measures of excess mortality, https://www.ons.gov.uk/peoplepopulationandcommunity/birthsdeathsandmarriages/deaths/articles/comparingdifferentinternationalmeasuresofexcessmortality/2022-12-20; 2022 [last accessed 8 April 2024].

6 Office for National Statistics. Death registration summary statistics, England and Wales: 2022, https://www.ons.gov.uk/peoplepopulationandcommunity/birthsdeathsandmarriages/deaths/articles/deathregistrationsummarystatisticsenglandandwales/2022#data-sources-and-quality; 2023 [last accessed 8 April 2024].

7 Islam N, Jdanov D A. Age and sex adjustments are critical when comparing death rates BMJ 2023; 381:p845 10.1136/bmj.p845.

8 Office for National Statistics. Estimating excess deaths in the UK, methodology changes, Estimating excess deaths in the UK, methodology changes - Office for National Statistics (ons.gov.uk); 2024 [last accessed 8 April 2024].

9 Fenton L, Minton J, Ramsay J, Kaye-Bardgett M, Fischbacher C, Wyper G M A, et al. Recent adverse mortality trends in Scotland: comparison with other high-income countries. BMJ Open 2019;9:e029936 10.1136/bmjopen-2019-029936.

10 Ho JY, Hendi AS. Recent trends in life expectancy across high income countries: retrospective observational study. BMJ 2018; 362: k2562 10.1136/bmj.k2562.

11 Public Health England. Recent trends in mortality in England: review and data packs. A report on recent trends in life expectancy and mortality in England. London, Public Health England, https://www.gov.uk/government/publications/recent-trends-in-mortality-in-england-review-and-data-packs; 2018 [last accessed 8 April 2024].

12 McCartney G, McMaster R, Popham F, Dundas R, Walsh D. Is austerity a cause of slower improvements in mortality in high-income countries? A panel analysis. Social Science and Medicine 2022; 313: 115397, 10.1016/j.socscimed.2022.115397.

13 Richardson E, McCartney G, Taulbut M, Douglas M, Craig N, et al. Population mortality impacts of the rising cost of living in Scotland: scenario modelling study. BMJ Public Health 2023;1:e000097. 10.1136/bmjph-2023-000097.

14 McCartney G, Douglas M, Taulbut M, Katikireddi SV, McKee M. Tackling population health challenges as we build back from the pandemic. BMJ 2021; 375: e066232, 10.1136/bmj-2021-066232.

15 Peters MDJ, Godfrey C, McInerney P, Munn Z, Tricco AC, Khalil, H. Chapter 11: Scoping Reviews (2020 version). In: Aromataris E, Munn Z, editors. JBI Manual for Evidence Synthesis, JBI, 2020, 10.46658/JBIMES-20-12.

16 Douglas M, McCartney G, Walsh D, Donaghy G, Rae D, Wild S, et al. Explanations for higher-than-expected mortality from April 2021: a scoping review protocol. medRxiv [pre-print]; 2023. 10.1101/2023.06.20.23291333.

17 Fadlallah R, Daher N, El-Harakeh A, Hammam R, Brax H, Bou L, et al. Approaches to prioritising primary health research: a scoping review. BMJ Global Health 2022;7:e007465. 10.1136/bmjgh-2021-007465.

18 Alagoz O, Lowry K P, Kurian A W, Mandelblatt J S, Ergun M A, Huang H, et al. Impact of disruptions in breast cancer control due to the COVID19 pandemic on breast cancer mortality in the United States: Estimates from collaborative simulation modeling. J Natl Cancer Inst. 2021;113(11):1484–1494. 10.1093/jnci/djab097.

19 Ali, S. ‘Huge, huge numbers:’ insurance group sees death rates up 40 percent over pre-pandemic levels, https://thehill.com/changing-america/well-being/longevity/588738-huge-huge-numbers-death-rates-up-40-percent-over-pre/; 2022 [last accessed 8 April 2024].

20 Alicandro G, Gerli A G, Remuzzi G, Centanni S, La Vecchia C. Updated estimates of excess total mortality in Italy during the circulation of the BA.2 and BA.4-5 Omicron variants: April-July 2022. La Medicina del lavoro 2022; 113(5):e2022046 10.23749/mdl.v113i5.13825.

21 Bilinski A, Thompson K, Emanuel E. COVID-19 and Excess All-Cause Mortality in the US and 20 Comparison Countries, June 2021-March 2022. JAMA 2023; 329(1):92–94. 10.1001/jama.2022.21795.

22 Bowden, M. Post-COVID excess mortality rates: What do they tell us about the state of public health in Europe? https://www.euronews.com/2023/03/11/excess-morality-rates-what-do-they-tell-us-about-the-state-of-public-health-in-europe; 2023 [last accessed 8 April 2024].

23 Buettner D, Hyman M. Why Is Our Life Expectancy Lower? #shorts. https://www.youtube.com/watch?v=J0a01mq7_GI; 2022 [last accessed 8 April 2024].

24 CDC National Center for Health Statistics. New Report Confirms U.S. Life Expectancy has Declined to Lowest Level Since 1996. https://www.cdc.gov/nchs/pressroom/nchs_press_releases/2022/20221222.htm; 2022 [last accessed 8 April 2024].

25 Coi G, Weise Z. Excess deaths surged as heat wave hit Europe. https://www.politico.eu/article/excess-death-surged-heat-wave-hit-europe/; 2022 [last accessed 8 April 2024].

26 Cuffe R, Schraer R. Excess deaths in 2022 among worst in 50 years, https://www.bbc.co.uk/news/health-64209221; 2023 [last accessed 8 April 2024].

27 Degeling K, Baxter N N, Emery J, Jenkins M A, Franchini F, Gibbs P, et al. An inverse stage-shift model to estimate the excess mortality and health economic impact of delayed access to cancer services due to the COVID-19 pandemic. Asia-Pacific Journal of Clinical Oncology 2021; 17(4):359–367. 10.1111/ajco.13505.

28 Department of Health and Social Care. Investigate UK excess deaths not related to Covid (petition response), https://petition.parliament.uk/petitions/628188#; 2023 [last accessed 8 April 2024].

29 DeVries A, Shambhu S, Sloop S, Overhage J M. One-Year Adverse Outcomes Among US Adults With Post-COVID-19 Condition vs Those Without COVID-19 in a Large Commercial Insurance Database. JAMA Health Forum 2023; 4(3):e230010. 10.1001/jamahealthforum.2023.0010.

30 Faguy A (2023). Extreme Heat in Southern Italy Increased July Death Rate, Officials Say, https://www.forbes.com/sites/anafaguy/2023/08/07/extreme-heat-in-southern-italy-increased-july-death-rate-officials-say/; 2023 [last accessed 8 April 2024].

31 Faust J S, Renton B, Chen A J, Du C, Liang C, Li S X, et al. Uncoupling of all-cause excess mortality from COVID-19 cases in a highly vaccinated state. The Lancet Infectious diseases 2022; 22(10):1419–1420. 10.1016/S1473-3099(22)00547-3.

32 Faust J S, Renton B, Du C, Chen A J, Li S-X, Lin Z, et al. Excess Mortality in the Vaccination Era in the United States, By State and 6-Month Period. medRxiv [pre-print]; 2023 10.1101/2023.03.07.23286927.

33 Ferenci T. Comparing methods to predict baseline mortality for excess mortality calculations - unravelling ‘the German puzzle’ and its implications for spline-regression. medRxiv [pre-print]; 2022 10.1101/2022.07.18.22277746.

34 Fouillet A, Martin D, Pontais I, Caserio-Schonemann C, Rey G. Reactive surveillance of suicides during the COVID-19 pandemic in France, 2020 to March 2022. Epidemiology and psychiatric sciences, 2023;32(101561091):e20. 10.1017/S2045796023000148.

35 Garver R (2022). US Life Expectancy Drops to Lowest in a Generation, https://www.voanews.com/a/us-life-expectancy-drops-to-lowest-in-a-generation/6889889.html; 2022 [last accessed 8 April 2024].

36 Han L, Zhao S, Li S, Gu S, Deng X, Yang L, et al. Excess cardiovascular mortality across multiple COVID-19 waves in the United States from March 2020 to March 2022. Nat Cardiovasc Res 2023; 2: 322–333. 10.1038/s44161-023-00220-2.

37 Harrison D. What is driving all cause excess mortality? BMJ 2022; 376: o100 10.1136/bmj.o100.

38 Health Feedback. What can explain the excess mortality in the U.S. and Europe in 2022?, https://healthfeedback.org/what-can-explain-the-excess-mortality-in-the-u-s-and-europe-in-2022/; 2022 [last accessed 8 April 2024].

39 Heuveline P. The Covid-19 pandemic and the expansion of the mortality gap between the United States and its European peers. PloS One 2023, 18(3):e0283153. 10.1371/journal.pone.0283153.

40 Hunter P. Summer 2022 saw thousands of excess deaths in England and Wales – here’s why that might be. https://theconversation.com/summer-2022-saw-thousands-of-excess-deaths-in-england-and-wales-heres-why-that-might-be-189351; 2022 [last accessed 8 April 2024].

41 Hussain Z. England and Wales have seen rise in excess deaths in 2022. BMJ 2022a; 378:o2283 10.1136/bmj.o2283.

42 Hussain Z. UK health officials analyse recent rise in excess deaths. BMJ 2022b;378:o2085. 10.1136/bmj.o2085.

43 Iacobucci G. England and Wales see rise in excess deaths amid flu surge. BMJ 2023; 380:p40, 10.1136/bmj.p40.

44 Ioannidis J P, Zonta F, Levitt M (2023). Variability in excess deaths across countries with different vulnerability during 2020-2023. medRxiv [pre-print]; 2023, 10.1101/2023.04.24.23289066.

45 Jones O. Britain’s excess death rate is at a disastrous high – and the causes go far beyond Covid, https://www.theguardian.com/commentisfree/2023/jan/15/britain-excess-death-rate-covid-nhs-cost-of-living; 2023 [last accessed 8 April 2024].

46 Jordan-Rios A, Nuzzi V, Bromage D I, McDonagh T, Sinagra G, Cannata A. Reshaping care in the aftermath of the pandemic. Implications for cardiology health systems. European journal of internal medicine 2023; 109: 4–11. 10.1016/j.ejim.2022.11.029.

47 Klobucista C (2022). U.S. Life Expectancy Is in Decline. Why Aren’t Other Countries Suffering the Same Problem? https://www.cfr.org/in-brief/us-life-expectancy-decline-why-arent-other-countries-suffering-same-problem; 2022 [last accessed 8 April 2024].

48 Konig S, Hohenstein S, Leiner J, Hindricks G, Meier-Hellmann A, Kuhlen R, et al. National mortality data for Germany before and throughout the pandemic: There is an excess mortality exceeding COVID-19-attributed fatalities. The Journal of infection 2022; 84(6):834–872. 10.1016/j.jinf.2022.02.024.

49 Lee W-E, Woo Park S, Weinberger D M, Olson D, Simonsen L, Grenfell B T, et al. Direct and indirect mortality impacts of the COVID-19 pandemic in the United States, March 1, 2020 to January 1, 2022. eLife 2023; 12: e77562 10.7554/eLife.77562.

50 Li S, Han L, Shi H, Chong M K, Zhao S, Ran J. Excess deaths from Alzheimer’s disease and Parkinson’s disease during the COVID-19 pandemic in the USA. Age and ageing 2022; 51 (12): afac277. 10.1093/ageing/afac277.

51 Liu J-Y, Chen T-J, Hou M-C (2021). Does COVID-19 vaccination cause excess deaths? Journal of the Chinese Medical Association 2021; 84(9):811–812. 10.1097/JCMA.0000000000000580.

52 Ludden I G, Jacobson S H, Jokela J A (2022). Excess deaths by sex and Age Group in the first two years of the COVID-19 pandemic in the United States. Health care management science 2022; 25(3):515–520. 10.1007/s10729-022-09606-3.

53 Lytras T, Athanasiadou M, Demetriou A, Stylianou D, Heraclides A, Kalakouta O (2022). Excess mortality in Cyprus during the COVID-19 pandemic and its lack of association with vaccination rates, medRxiv [pre-print]; 2022, 10.1101/2022.08.05.22278487.

54 Mattiuzzi C, Henry B M, Lippi G (2023). Correlation between relative age-standardized mortality rates and COVID-19 mortality over time in Italy. Acta Biomed 2023; 94(2):e2023056. 10.23750/abm.v94i2.14139.

55 McDonough S. Why are American lives getting shorter? Retrieved from Vox: https://www.vox.com/future-perfect/2022/9/7/23339734/life-expectancy-shorter-united-states-covid; 2022 [last accessed 8 April 2024].

56 Ministry of Health Singapore (2022). Report on excess mortality during the COVID-19 pandemic up to June 2022. https://www.moh.gov.sg/docs/librariesprovider5/resources-statistics/reports/report-on-excess-mortality-during-the-covid-pandemic-18sep2022.pdf; 2022 [last accessed 8 April 2024].

57 Motairek I, Janus S E, Hajjari J, Nasir K, Khan S U, Rajagopalan S, et al. Social Vulnerability and Excess Mortality in the COVID-19 Era. The American journal of cardiology 2022; 172: 172–174. 10.1016/j.amjcard.2022.03.011.

58 Mulligan C, Arnott R (2022). Non-Covid Excess Deaths, 2020-21: Collateral Damage of Policy Choices?, https://www.nber.org/papers/w30104; 2022 [ast accessed 8 April 2024].

59 Nakanishi M, Yamasaki S, Endo K, Ando S, Sakai M, Yoshii H, et al. Suicide rates during the COVID-19 pandemic in Japan from April 2020 to December 2021. Psychiatry research 2022; 316: 114774. 385):114774. 10.1016/j.psychres.2022.

60 Odd D, Stoianova S, Williams T, Fleming P, Luyt K. Child Mortality in England During the First 2 Years of the COVID-19 Pandemic. JAMA network open 2023; 6(1):e224919. 10.1001/jamanetworkopen.2022.49191.

61 Orchard J, Workman C, Puranik R. Endemic Covid is contributing to significant excess cardiac mortality. J Am Coll Cardiol 2023; 81: (8_Supplement) 2361. 10.1016/S0735-1097(23)02805-X.

62 Paglino E, Lundberg D J, Zhou Z, Wasserman J A, Raquib R, Luck A N, et al. Monthly excess mortality across counties in the United States during the COVID-19 pandemic, March 2020 to February 2022. Sci Adv 2023a; 9(25):eadf9742. 10.1126/sciadv.adf9742.

63 Paglino E, Lundberg D J, Zhou Z, Wasserman J A, Raquib R, Hempstead K, et al. (2023b). Differences Between Reported COVID-19 Deaths and Estimated Excess Deaths in Counties Across the United States, March 2020 to February 2022. medRxiv [pre-print]; 2023b, 10.1101/2023.01.16.23284633.

64 Ricoca Peixoto V, Vieira A, Aguiar P, Abrantes A. Excess Mortality since COVID-19: What Data Do We Need and What Questions Should We Ask to Understand its Causes in Portugal? Acta Médica Portuguesa 2022; 35(11):783–785, 10.20344/amp.19080.

65 Riou J, Hauser A, Fesser A, Althaus C, Egger M, Konstantinoudis G. (2022). Direct and indirect effects of the COVID-19 pandemic on mortality in Switzerland: A population-based study. medRxiv [pre-print]; 2022, 10.1101/2022.08.05.22278458.

66 Roccetti M. Excess mortality and COVID-19 deaths in Italy: A peak comparison study. Mathematical biosciences and engineering 2023; 20(4):7042–7055. 10.3934/mbe.2023304.

67 Rosen M, Stenbeck M. Interventions to suppress the coronavirus pandemic will increase unemployment and lead to many premature deaths. Scandinavian journal of public health 2021; 49(1):64–68. 10.1177/1403494820947974.

68 Seheult R. Excess Mortality and COVID-19: What is causing it?, https://www.youtube.com/watch?v=vMC9dvbR82g; 2022 [last accessed 8 April 2024].

69 Seheult R. Excess Deaths: Causes (Deep Dive), https://www.youtube.com/watch?v=-ZI7dwldSR0; 2023 [last accessed 8 April 2024].

70 Sforza L. Misinformation contributing to lower life expectancy in US, FDA chief says, https://thehill.com/policy/healthcare/3944280-misinformation-contributing-to-lower-life-expectancy-in-us-fda-chief-says/; 2023 [last accessed 8 April 2024].

71 Smart D, Riccelli P, Kerr K, Clark J, Fosvig S, Lawler M. Significant decline in cancer diagnostic testing in U.S. CMS population during the COVID-19 pandemic. Journal of Clinical Oncology 2021; 39(15), supplement. 10.1200/JCO.2021.39.15_suppl.653.

72 Šorli A S, Makovec T, Krevel Ž, Gorjup R. Forgotten "Primum Non Nocere" and Increased Mortality after Covid-19 Vaccination. Quality in Primary Care 2023; 31(1); 15–34. 10.36648/1479-1064.23.31.06.

73 Statbel. 5.5% more deaths in 2022, https://statbel.fgov.be/en/news/55-more-deaths-2022; 2023 [last accessed 8 April 2024].

74 Stoto M A, Schlageter S, degl’Innocenti D G, Zollo F, Kraemer J D. Differential COVID-19 mortality in the United States: Patterns, causes and policy implications. medRxiv [pre-print]; 2023, 10.1101/2023.01.09.23284358.

75 The King’s Fund. The King’s Fund responds to latest ONS comparisons of all-cause mortality between European countries during the pandemic, https://www.kingsfund.org.uk/press/press-releases/kings-fund-responds-latest-ons-comparisons-all-cause-mortality-between; 2022 [last accessed 8 April 2024].

76 The King’s Fund. The King’s Fund responds to the latest ONS mortality data, https://www.kingsfund.org.uk/press/press-releases/december%202021-latest-ons-mortality-data; 2023 [last accessed 8 April 2024].

77 Todd M, Scheeres A. Excess Mortality From Non-COVID-19 Causes During the COVID-19 Pandemic in Philadelphia, Pennsylvania, 2020-2021. American journal of public health 2022; 112(12):1800–1803. 10.2105/AJPH.2022.307096.

78 UKHSA. UKHSA and ONS release estimates of excess deaths during summer of 2022, https://www.gov.uk/government/news/ukhsa-and-ons-release-estimates-of-excess-deaths-during-summer-of-2022; 2022 [last accessed 8 April 2024].

79 van den Puttelaar R, Landsdorp-Vogelaar I, Hahn A I, Rutter C M Levin T R, Zauber A G, et al. Impact and Recovery from COVID-19-Related Disruptions in Colorectal Cancer Screening and Care in the US: A Scenario Analysis. Cancer Epidemiol Biomarkers Prev 2023; 32 (1): 22–29. 10.1158/1055-9965.EPI-22-0544.

80 Wedler M, Pinto J G, Hochman A. More frequent, persistent, and deadly heat waves in the 21st century over the Eastern Mediterranean. The Science of the total environment 2023; 870: 161883. 10.1016/j.scitotenv.2023.161883.

81 Woolf S H. Excess Deaths Will Continue In The United States Until The Root Causes Are Addressed. Health affairs 2022; 41(11):1562–1564. 10.1377/hlthaff.2022.01103.

82 Woolf S H, Wolf E R, Rivara F P. The New Crisis of Increasing All-Cause Mortality in US Children and Adolescents. JAMA 2023; 329(12):975–976. 10.1001/jama.2023.3517.

83 Yao X I, Han L, Sun Y, He D, Zhao S, Ran J. Temporal variation of excess deaths from diabetes during the COVID-19 pandemic in the United States. Journal of infection and public health 2023; 16(4):483–489. 10.1016/j.jiph.2023.01.018.

84 Yeo Y H, Wang M, He X, Lv F, Zhang Y, Zu J, et al. Excess risk for acute myocardial infarction mortality during the COVID-19 pandemic. Journal of medical virology 2023; 95(1):e28187. 10.1002/jmv.28187.

85 Ylli A, Burazeri G, Wu Y Y, Sentell T. COVID-19 excess death rate in Eastern European countries associated with weaker regulation implementation and lower vaccination coverage. medRxiv [pre-print]; 2022, 10.1101/2022.02.06.22270549.

86 Yoshioka E, Hanley S J, Sato Y, Saijo Y. Impact of the COVID-19 pandemic on suicide rates in Japan through December 2021: An interrupted time series analysis. The Lancet regional health. Western Pacific 2022; 24:100480. 10.1016/j.lanwpc.2022.100480.

87 Page M, McKenzie J E, Bossuyt P M, Boutron I, Hoffman T C, Mulrow C D, et al. The PRISMA 2020 statement: an updated guideline for reporting systematic reviews. BMJ, 372: n71 10.1136/bmj.n71.

88 Hiam L, Dorling D, McKee M. When experts disagree: interviews with public health experts on health outcomes in the UK 2010-2020. Public Health 2023; 214:96–105. 10.1016/j.puhe.2022.10.019.

89 Whitty C J M, Smith G, McBride M, Atherton F, Powis S H, Stokes-Lampard H. Restoring and extending secondary prevention. BMJ 2023; 380: 201 10.1136/bmj.p201.

90 Walsh D, McCartney G, Minton J, Parkinson J, Shipton D, Whyte B Changing mortality trends in countries and cities of the UK: a population-based trend analysis. BMJ Open 2020; 10: e038135 10.1136/bmjopen-2020-038135.

91 McCartney G, Walsh D, Fenton L, Devine R. Resetting the course for population health: evidence and recommendations to address stalled mortality improvements in Scotland and the rest of the UK. Glasgow: Glasgow Centre for Population Health/University of Glasgow; 2022.

92 ^92^ Bradford D R R, Brown D, McCartney G, Douglas M, Dundas R, Walsh D. Post-pandemic excess mortality in England and Scotland: is it a continuation of pre-pandemic trends? Conference abstract (submitted to Society of Social Medicine & Population Health conference 2024).

93 Walsh D, Dundas R, McCartney G, Gibson M, Seaman R. Bearing the burden of austerity: how do changing mortality rates in the UK compare between men and women? Journal of Epidemiology & Community Health 2022; 76: 1027–1033. 10.1136/jech-2022-219645.

94 National Records of Scotland (NRS). COVID-19 deaths 2022. Edinburgh: NRS. https://www.nrscotland.gov.uk/files//statistics/covid19/covid-19-annual-deaths-2022-report.pdf; 2023 [last accessed 8 April 2024].

95 Shah S A, Robertson C, Sheikh A. Effects of the COVID-19 pandemic on NHS England waiting times for elective hospital care: a modelling study. Lancet 2024; 403 (10423): 241–243. 10.1016/S0140-6736(23)02744-7.

96 Douglas M, Katikireddi S V, Taulbut M, McKee M, McCartney G. Mitigating the wider health effects of covid-19 pandemic response. BMJ 2020; 369: m1557. 10.1136/bmj.m1557.

97 Marmot M, Allen J, Goldblatt P, Herd E, Morrison J. Build Back Fairer: The COVID-19 Marmot Review. The Pandemic, Socioeconomic and Health Inequalities in England. London: Institute of Health Equity; 2023.

